# Risk Factor Analysis for Extended-Spectrum Beta-Lactamase Producing Enterobacterales Colonization or Infection: Evaluation of a Novel Approach to Assess Local Prevalence as a Risk Factor

**DOI:** 10.1101/2022.12.22.22283612

**Authors:** Jerald Cherian, Sara E. Cosgrove, Fardad Haghpanah, Eili Y. Klein, the Centers for Disease Control and Prevention’s Prevention Epicenters Program and the Modeling Infectious Diseases in Healthcare Network

## Abstract

**Objective:** To explore an approach to identify the risk of local prevalence of extended-spectrum beta-lactamase producing Enterobacterales (ESBL-E) on ESBL-E colonization or infection, and reassess known risk factors.

**Design:** Case-control study.

**Setting:** Johns Hopkins Health System emergency departments (EDs) in the Baltimore-Washington, D.C. region.

**Patients:** Patients aged ≥18 years with a culture growing Enterobacterales between 4/2019-12/2021. Cases had a culture growing an ESBL-E.

**Methods:** Addresses were linked to Census Block Groups and placed into communities using a clustering algorithm. Prevalence in each community was estimated by the proportion of ESBL-E among Enterobacterales isolates. Logistic regression was used to determine risk factors for ESBL-E colonization or infection.

**Results:** ESBL-E were detected in 1167 of 11,229 patients (10.4%). Risk factors included a history of ESBL-E in the prior year (OR, 15.62; 95% CI, 11.25-21.68), and exposure to a skilled nursing or long term care facility (OR, 1.66; 95% CI, 1.39-1.99), 3rd generation cephalosporin (OR, 1.83; 95% CI, 1.50-2.23), carbapenem (OR, 2.01; 95% CI, 1.46-2.78) or trimethoprim-sulfamethoxazole (OR, 1.53; 95% CI, 1.05-2.22) within the prior 6 months. Patients were at lower risk if their community had a prevalence <25^th^ percentile in the prior 3 months (OR, 0.84; 95% CI, 0.71-0.98), 6 months (OR, 0.84; 95% CI, 0.71-0.99) or 12 months (OR, 0.81; 95% CI, 0.69-0.96). There was no association between being in a community >75^th^ percentile and the outcome.

**Conclusions:** This method of defining the recent local prevalence of ESBL-E may partially capture differences in the likelihood of a patient having an ESBL-E.

## Introduction

Infections due to ESBL-E are associated with increased morbidity and mortality compared to infections due to non-ESBL-E.^1,2^ Prior data suggests that this is in part due to delays in the initiation of effective antimicrobial therapy.^3,4^ ESBL-E are resistant to many commonly used beta-lactam antibiotics and may also harbor resistance to other classes of antibiotics but remain susceptible to carbapenems.^5,6^ However, judicious carbapenem use in clinical practice is essential to reduce the risk of selecting for carbapenem-resistant organisms.^7^ There is a need to further understand risk factors for infection with ESBL-Es, particularly among patients presenting to the ED where results of rapid diagnostic tests are typically unavailable at the time of empiric antibiotic selection, to inform antibiotic decision-making.

Recent residence in or travel to a country with a high prevalence of ESBL-E has been identified as an ESBL-E risk factor.^8,9^ The prevalence of ESBL-E not only varies at the country level, but also within smaller geographic regions such as a county or city.^10,11^ These differences in prevalence are due to specific community characteristics that affect the transmission dynamics of ESBL-E and place patients in distinct communities at different risk for ESBL-E colonization or infection.^12–17^ Identifying whether a patient resides in a community with high ESBL-E prevalence can help clinicians risk stratify patients. However, no studies have investigated how to distinguish these different local communities in order to estimate the risk of ESBL-E infection or colonization at the patient level due to the community prevalence of ESBL-E. U.S. Census Block Group (CBG) data can be used to cluster patients into aggregates based on shared demographics and geography, which may approximate different communities in a geographic region that have distinct transmission dynamics and prevalence of ESBL-E.

This study had two objectives: 1) to reassess previously identified risk factors for ESBL-E colonization or infection among ED patients, and 2) to assess whether CBG data can be used to cluster patients into local geographic aggregates that approximate local communities with differences in ESBL-E transmission to more accurately capture a patient’s risk of ESBL-E colonization or infection based on the local prevalence in the patient’s area of residence.

## Methods

### Study Setting and Population

This study included patients aged ≥18 years who presented to Johns Hopkins Health System (JHHS) EDs located in the Baltimore-Washington, D.C. region (Johns Hopkins Hospital and Johns Hopkins Bayview Medical Center, Baltimore, Maryland; Howard County General Hospital, Columbia, Maryland; Suburban Hospital, Bethesda, Maryland; and Sibley Memorial Hospital, Washington, D.C.) from 4/2019 to 12/2021 with a culture obtained in the ED that grew *Escherichia coli, Klebsiella pnuemoniae, Klebsiella oxytoca, or Proteus mirabilis*. Only these organisms were included because 1) they are the most common organisms that produce ESBLs and 2) the microbiology laboratory employed phenotypic testing to confirm ESBL status for isolates. Patients with these organisms identified via stool surveillance cultures were excluded. Case patients had at least one culture obtained in the ED that grew an ESBL-producing isolate while control patients did not have an ESBL-producing isolate identified. Only the most recent ED encounter for a given patient was included. This study was approved by the Johns Hopkins University School of Medicine Institutional Review Board with a waiver of informed consent.

### Data Collection

Clinical data were extracted from the time of presentation to the ED from a limited dataset of JHHS ED patients utilizing the Johns Hopkins Precision Medicine Analytics Platform, that includes data from the JHHS electronic health record system (Epic). Data included demographics, preexisting medical conditions, and presence of devices at presentation; hospitalizations, intensive care unit admissions and microbiologic data in the previous year; and long-term care facility or nursing home admissions, procedures, and antibiotics and gastric acid suppressant use in the previous 6 months. When assessing for a prior history of ESBL-E in the past year, only *E. coli, K. pneumoniae, K. oxytoca, or P. mirabilis* isolates were included. Data regarding recent international travel, clinical symptoms, and specific ESBL genes in the ESBL-positive organisms were not available.

### Microbiology Methods

Bacterial cultures were processed at a JHHS clinical microbiology laboratory according to standard operating procedures. Blood cultures growing a Gram-negative organism underwent testing with the GenMark Dx ePlex blood culture identification Gram-negative (BCID-GN) panel. BCID-GN testing was limited to the first positive blood culture for a patient. Detection of *bla*_CTX-M_ genes on the BCID-GN panel was considered confirmatory for ESBL status.

Additionally, all *E. coli, K. pnuemoniae, and K. oxytoca* underwent automated screening and confirmation of ESBL status by the BD Phoenix Automated System (BD Diagnostics, Sparks, Maryland). All *P. mirabilis* isolates with a ceftriaxone or ceftazidime minimum inhibitory concentration (MIC) ≥2 μg/mL underwent additional disk diffusion testing using both cefotaxime and ceftazidime, alone and in combination with clavulanate. A ≥5mm increase in the zone of diameter for either agent tested in combination with clavulanate compared to when tested alone was considered confirmatory for ESBL status.

### Geospatial Analysis

The assumption underlying the geospatial analysis was that ESBL-Es are more likely to transmit between individuals that are part of a community than to people outside that community. To define communities, we used data from SafeGraph (www.safegraph.com), which aggregates anonymized location data from mobile devices to provide insights about movement patterns, to create clusters of CBGs that are more connected. Data on movement for the CBGs in the Baltimore-Washington, D.C. region were extracted from SafeGraph from January 2019 to March 2021. We used the greedy Clauset-Newman-Moore^18^ and the Louvain algorithm^19^ community detection algorithms to discover clusters of CBGs in which residents are more likely to interact with each other rather than with people from other clusters (see Supplementary information for complete methods).

Alteryx (www.alteryx.com), a software platform for spatial data analytics, was used to retrieve census block group information based on the patient home address. Patient CBGs were matched to community clusters and the prevalence of ESBL-E in the prior three, six and twelve months was calculated using the number of ESBL-E out of the total number of *E. coli, K. pnuemoniae, K. oxytoca, or P. mirabilis* isolates within each patient’s community. As the community detection algorithm did not assign every CBG to a community, if a patient was not included in a community, the median prevalence of ESBL-E among all communities was imputed for the 3 month, 6 month, and 12 month prevalence of ESBL-E in their community. For each patient the prevalence of ESBL-E in the period 3 months, 6 months, and 12 months prior was categorized as being in the <25^th^ percentile, between the 25^th^ and 75^h^ percentile, or above the 75^th^ percentile among all communities during the respective period.

### Statistical Analysis

Descriptive statistics for patient variables were calculated using median (interquartile range) or frequency count (percentage) as appropriate. Univariable logistic regression models were used to assess the relationship between potential risk factors identified in the prior literature and ESBL status. A multivariable logistic regression model was then constructed using the same potential risk factors used in univariable analyses (Model 1). Given clinical significance and evidence of association with the outcome in prior literature, all variables were retained in the model even if they were not statistically significant. Additional multivariable models were constructed using the same variables with the addition of a categorical variable indicating the percentile for the community’s ESBL-E prevalence in the past 3 months (Model 2), 6 months (Model 3), or 12 months (Model 4) prior to presentation. Models 2, 3 and 4 were compared to Model 1 using likelihood-ratio tests. The goodness of fit was assessed using the Hosmer-Lemeshow test. The variable inflation factor was used to assess for collinearity. Subgroup analysis was also performed including only patients without a history of an ESBL-positive culture in the prior year utilizing the same multivariable models to assess for potential effect modification based on history of ESBL positive culture. All significance testing was done at an α level of 0.05. Data were analyzed using STATA version 17.0 (StataCorp, College Station, TX).

## Results

From April 2019 to December 2021, there were a total of 12,905 bacterial cultures from 11,229 patients that grew *E. coli* (68.8%), *K. pnuemoniae* (18.7%), *K. oxytoca* (2.9%), *or P. mirabilis* (9.6%). Among all organisms, 16.1% were isolated from blood, 78.3% from urine, 0.3% from a respiratory source, and 5.3% from other sources. A total of 1,167 patients (10.4%) had at least one culture growing an ESBL-E. The median age of patients was 69 years (IQR, 49–82), 29.0% were male, and 53.3% were non-Hispanic white (Table 1). Sixty-five percent of patients were admitted to the hospital. In the preceding year, 259 patients (2.3%) had a prior culture that grew an ESBL-E. In the preceding 6 months, 26.3% of patients had received at least one dose of antibiotics and 14.1% had exposure to a skilled nursing facility or long-term care facility. In the previous 12 months, 23.5% had been hospitalized, 4.9% had been admitted to an intensive care unit (ICU), and 18.1% had a surgery or procedure.

**Table 1.** Baseline Characteristics for Emergency Department Patients with *Escherichia coli, Klebsiella pnuemoniae, Klebsiella oxytoca, or Proteus mirabilis* Isolated in Culture, by Extended-Spectrum β-Lactamase Status

| Baseline Characteristics | Total Cohort<br>(N=11229) | ESBL Positive<br>(N=1167) | ESBL Negative<br>(N=10062) |
| --- | --- | --- | --- |
| Age group – No. (%) |  |  |  |
| 18-25 yr | 647 (5.8) | 28 (2.4) | 619 (6.2) |
| 26-35 yr | 990 (8.8) | 89 (7.6) | 901 (9.0) |
| 36-45 yr | 858 (7.6) | 68 (5.8) | 790 (7.9) |
| 46-55 yr | 1014 (9.0) | 113 (9.7) | 901 (8.6) |
| 56-65 yr | 1569 (14.0) | 198 (17.0) | 1371 (13.6) |
| 66-75 yr | 2010 (17.9) | 263 (22.5) | 1747 (17.4) |
| 76-85 yr | 2247 (20.0) | 253 (21.7) | 1994 (19.8) |
| >85 yr | 1894 (16.8) | 155 (13.3) | 1739 (17.3) |
| Male Sex – No. (%) | 3377 (29.0) | 705 (39.6) | 7147 (30.1) |
| Race/Ethnicity – No. (%) |  |  |  |
| White | 5984 (53.4) | 537 (46.0) | 5447 (54.1) |
| Black | 3267 (29.1) | 348 (29.8) | 2919 (29.0) |
| Asian | 573 (5.1) | 78 (6.7) | 495 (4.9) |
| Hispanic | 1000 (8.9) | 135 (11.6) | 865 (8.6) |
| Other | 405 (3.6) | 69 (5.9) | 336 (3.3) |
| Comorbidities – No. (%) |  |  |  |
| Diabetes | 2214 (19.7) | 323 (27.7) | 1891 (18.8) |
| Chronic Kidney Disease | 1598 (14.2) | 233 (20.0) | 1365 (13.6) |
| Chronic Pulmonary Disease | 1222 (10.9) | 153 (13.1) | 1069 (10.6) |
| Chronic Liver Disease | 477 (4.3) | 51 (4.4) | 426 (4.2) |
| Cancer | 1572 (14.0) | 165 (14.1) | 1407 (14.0) |
| HIV Positive | 1126 (10.0) | 167 (14.3) | 959 (9.5) |
| Antibiotic Use (<6 Months) – No. (%) |  |  |  |
| 3<sup>rd</sup> Generation Cephalosporin | 1314 (11.7) | 302 (25.9) | 1012 (10.1) |
| 4<sup>th</sup> Generation Cephalosporin | 608 (5.4) | 139 (11.9) | 469 (4.7) |
| Piperacillin-Tazobactam | 611 (5.4) | 122 (10.5) | 489 (4.9) |
| Carbapenem | 363 (3.2) | 167 (14.3) | 196 (2.0) |
| Fluoroquinolone | 389 (3.5) | 82 (7.0) | 307 (3.1) |
| Aminoglycoside | 111 (1.0) | 23 (2.0) | 88 (0.9) |
| Trimethoprim-Sulfamethoxazole | 239 (2.1) | 60 (5.1) | 179 (1.8) |
| Aztreonam | 51 (0.45) | 12 (1.0) | 39 (0.4) |
| Metronidazole | 472 (4.2) | 81 (6.9) | 391 (3.9) |
| Other Antibiotic | 1898 (16.9) | 307 (26.3) | 1591 (15.8) |
| Any Antibiotic | 2956 (26.3) | 537 (46.0) | 2419 (24.0) |
| Acid Suppressant Use (<6 Months) – No. (%) |  |  |  |
| Proton Pump Inhibitor | 1363 (12.1) | 247 (21.2) | 1116 (11.1) |
| H2 Antagonist | 717 (6.4) | 125 (10.7) | 592 (5.9) |
| Device or Hardware at Presentation |  |  |  |
| Urinary Catheter | 316 (2.8) | 66 (5.7) | 250 (2.5) |
| Tracheostomy | 29 (0.3) | 14 (1.2) | 15 (0.2) |
| Gastrointestinal Feeding Tube | 41 (0.4) | 9 (0.8) | 32 (0.3) |
| Central Line | 202 (1.8) | 32 (2.7) | 170 (1.7) |
| Healthcare Exposure – No. (%) |  |  |  |
| Hospitalization (<12 months) | 2640 (23.5) | 432 (37.0) | 2208 (21.9) |
| ICU Admission (<12 months) | 545 (4.9) | 121 (10.4) | 424 (4.2) |
| Surgery or Procedure (<12 months) | 2029 (18.1) | 296 (25.4) | 1733 (17.2) |
| Long Term Care Facility or Skill Nursing Facility Admission (<6 Months) | 1591 (14.2) | 2 88 (24.7) | 1303 (13.0) |
| History of ESBL-E (<12 months)– No. (%) | 259 (2.3) | 191 (16.4) | 68 (0.7) |
| Percent of ESBL-E Isolates within Patient’s CBG Community – Median (IQR) |  |  |  |
| <3 Months | 9.7 (8.3–11.1) | 9.7 (9.2–11.1) | 9.7 (8.2–11.2) |
| <6 Months | 10.3 (8.9–11.7) | 10.3 (9.5–11.5) | 10.3 (8.9–11.7) |
| <12 Months | 10.1 (9.3–11.0) | 10.1 (9.6–11.0) | 10.1 (9.2–11.0) |

Abbreviations: CBG = Census block group; ESBL-E = Extended-spectrum $\beta$ -lactamase producing Enterobacterales; H-2 = Histamine H2-receptor; HIV = Human Immunodeficiency Virus; ICU = Intensive care unit; OR = Odds Ratio

Among all CBG communities, the median prevalence of ESBL-E was 9.7% (IQR, 8.3%–11.1%) in the 3 months prior to presentation. The median percentage was 10.3% (IQR, 8.9%–11.7%) in the previous 6 months and 10.1% (IQR, 9.3%–11.0%) in the previous 12 months.

On univariable logistic regression analysis, the variables most strongly associated with the outcome were carbapenem use in the past 6 months (OR, 8.41; 95% CI, 6.77-10.43), presence of a tracheostomy on presentation (OR, 8.13; 95% CI, 3.92-16.89), and history of an ESBL-positive culture in the last year (OR, 28.76; 95% CI, 21.64-38.23) (Table S1). Patients had a lower odds of having an ESBL-E isolated in culture if they were in a community with an ESBL-E prevalence less than the 25^th^ percentile in the 3 months (OR, 0.84; 95% CI, 0.72-0.98) and 12 months (OR, 0.84; 95% CI, 0.72-0.98) prior to admission compared to patients in communities with an ESBL-E prevenance in the 25^th^ to 50^th^ percentile during those periods. This association was not significant for communities with ESBL-E prevalence less than the 25^th^ percentile in the 6 months prior to admission (OR, 0.86; 95% CI, 0.74-1.01). There was no significant association with the outcome for patients who were in communities with greater than the 75^th^ percentile for ESBL-E prevalence regardless of timeframe prior to presentation in which prevalence was assessed.

On multivariable analysis excluding the geospatial analysis, the odds of having an ESBL-E isolated in culture was higher among those who were 66-75 years of age (OR, 3.07; 95% CI, 2.00-4.74) and male (OR, 1.24; 95% CI, 1.08-1.43) (Table 2). Non-white patients were at higher risk with the highest odds being in Hispanic patients (OR, 2.22; 95% CI, 1.77-2.79) and patients who do not identify as white, black, Asian, or Hispanic (OR, 2.51; 95% CI, 1.86-3.39). A prior history of an ESBL-positive culture in the prior 12 months was strongly associated with ESBL-E isolation (OR, 15.62; 95% CI, 11.25-21.68). Exposure to a 3^rd^ generation cephalosporin (OR, 1.83; 95% CI, 1.50-2.23), carbapenem (OR, 2.01; 95% CI, 1.46-2.78), or trimethoprim-sulfamethoxazole (OR, 1.53; 95% CI, 1.05-2.22) in the 6 months prior to presentation were also independently associated with ESBL-E isolation. Patients with a tracheostomy (OR, 4.09, 95% CI, 1.74-9.60) and long-term care facility or skilled nursing facility exposure in the previous 6 months (OR, 1.66; 95% CI, 1.39-1.99) were at increased risk. A history of cancer (OR, 10.77; 95% CI, 0.63-0.94) was associated with a lower odds of ESBL-E isolation.

**Table 2.**
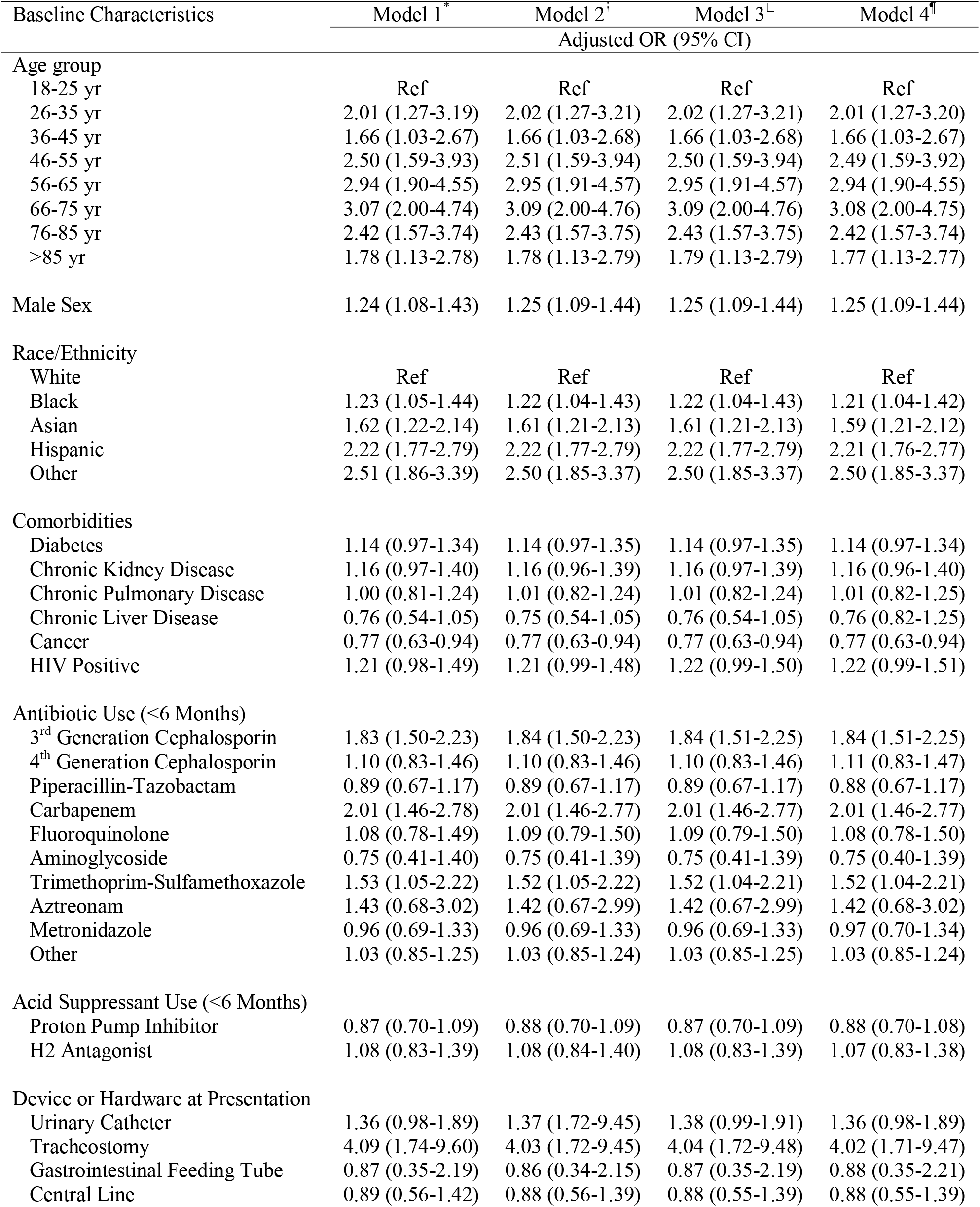
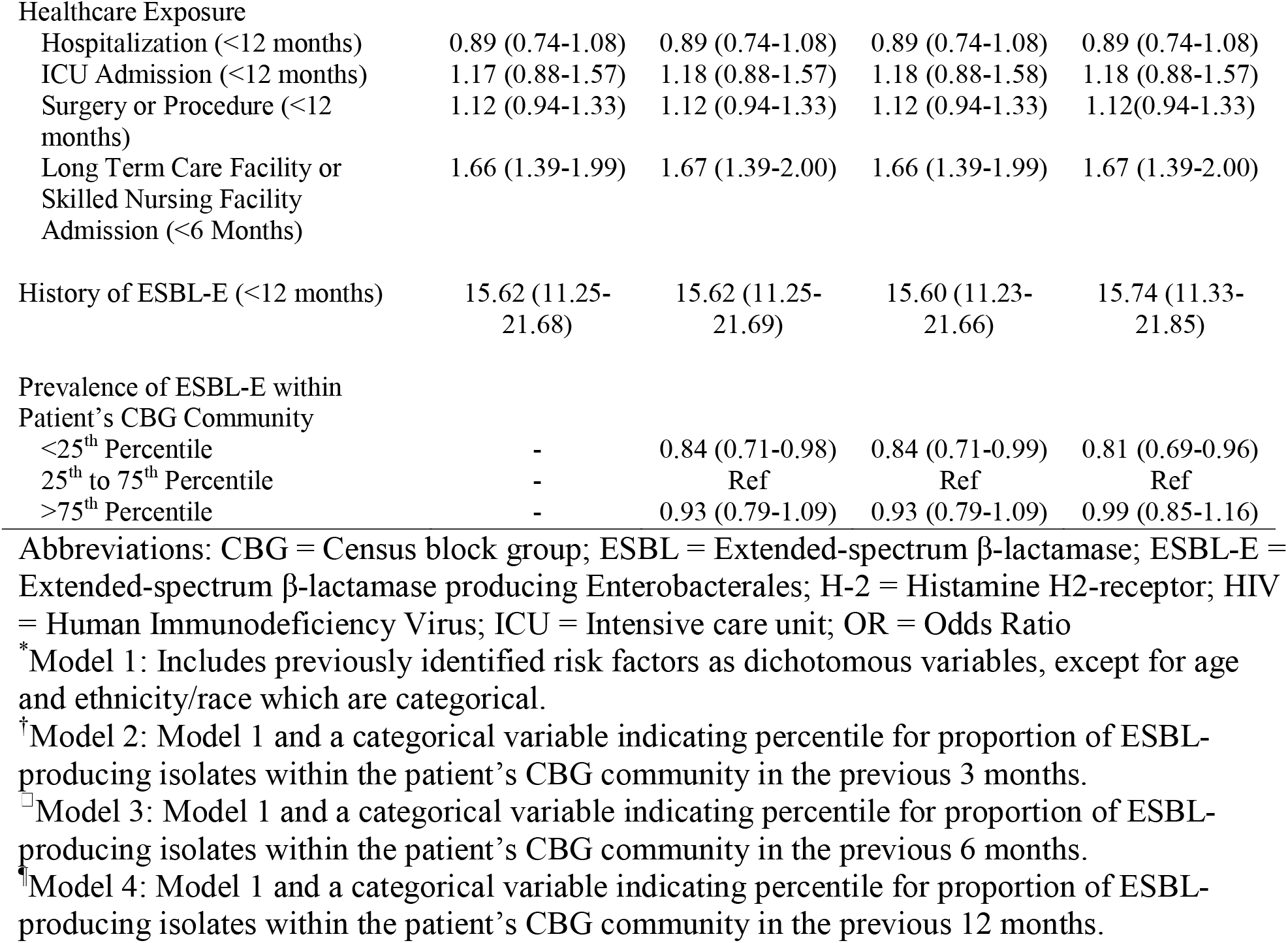
Multivariable Logistic Regression Models Evaluating Risk Factors for Isolation of an Extended-Spectrum β-Lactamase Producing Enterobacterales from Emergency Department Patients

There were no substantial differences in the adjusted odds ratios and corresponding confidence intervals for the variables when adding the geospatial analysis (Table 2). Patients in communities with an ESBL-E prevalence less than the 25^th^ percentile had a lower risk of having an ESBL-E isolated in culture compared to patients who were in communities with ESBL-E prevalence in the 25^th^ to 50^th^ percentile regardless of whether the prevalence was assessed in the prior 3 months (OR, 0.84; 95% CI, 0.71-0.98), 6 months (OR, 0.84; 95% CI, 0.71-0.99) or 12 months (OR, 0.81; 95% CI, 0.69-0.96). There was no significant association with the outcome for patients who were in communities with greater than the 75^th^ percentile for ESBL-E prevalence regardless of timeframe prior to presentation in which prevalence was assessed. Results of the multivariable regression models were qualitatively similar when including only patients without a history of an ESBL-producing organism in the past year (Table S2).

There was no evidence of collinearity and all multivariable models showed good fit. There was no improvement in model fit for the geospatial models when compared to the base model.

## Discussion

This study confirms the importance of several previously identified risk factors for ESBL-E colonization or infection, and demonstrates a weak association between the estimated local prevalence of ESBL-E using CBG aggregations and a patient’s risk of ESBL-E infection or colonization.

Similar to prior studies, the most significant risk factor was a recent history of an ESBL-E positive culture with 15.62 times higher odds of having an ESBL-E colonization or infection.^20,21^ Additionally, recent exposure to a long-term care or skilled nursing facility, 3^rd^ generation cephalosporins, and trimethoprim-sulfamethoxazole were associated with an increased risk.^20,22–25^ Recent carbapenem use was also associated with an increased risk even when controlling for a history of an ESBL-E in the past year. Although another study found the same association, the reason is uncertain.^22^ It may be due to residual confounding from providers accounting for more remote history of ESBL-positive cultures when prescribing antibiotics. Notably, patients with a history of cancer had a lower risk of having an ESBL-E isolated on culture, contrary to prior studies, perhaps due to behavioral factors that are more common in this group (e.g., social distancing) that reduce exposure to ESBL-E in the community.^25–27^

Community transmission of ESBL-Es has increased 8-fold over the past two decades globally.^28^ Patients who are from countries with higher prevalence are more likely to be colonized and colonization confers higher risk for development of an ESBL-E infection.^28–32^ Although differences in prevalence have most often been described on the country level, geospatial mapping has shown differences in prevalence within areas of a single city.^11^ The differences in prevalence within these local communities has been postulated to be related to specific environmental factors, such as contaminated food or water sources, presence of farmland with animals that harbor ESBL-E, or overcrowded housing that promotes transmission.^14–17^ Differences in the demographics also contribute, such a larger proportion of foreign-born residents leading to importation of ESBL-E from other countries into that local community.^12,13^ Additionally, the interconnectivity between individuals within an area likely plays a key role in transmission. Such factors lead to unique transmission dynamics within that specific local community and thus lead to a different prevalence of ESBL-E. Being able to identify if a patient resides in a local community with low or high ESBL-E prevalence would assist clinicians in assessing risk of ESBL-E infection for that patient. However, it is challenging to define these local communities given they are not defined by geography alone.

We used a novel approach to estimate local prevalence of ESBL-E by utilizing CBG data to cluster patients into aggregates based on geography and shared demographics to approximate the local communities in which there may be distinct ESBL-E transmission. Patients who were in communities that were below the 25^th^ percentile for prevalence had 16-19% lower odds of ESBL-E colonization or infection compared to those in the 25^th^ to 75^th^ percentile, although this is a weak association. There was no increased risk for patient’s in communities that were above the 75^th^ percentile in prevalence. The findings suggest that the methods used to aggregate CBGs may capture only some of the relationship between ESBL-E prevalence in local communities and the risk of ESBL-E colonization or infection. A method that accurately captures the variations in local prevalence and can inform risk stratification should show an increased risk of ESBL-E infection or colonization among patients in high prevalence areas and decreased risk for those in low prevalence areas. Further exploration of how geographic data can be used to aggregate patients into meaningful groups where transmission differs is warranted.

This study has some limitations. The majority of Enterobacterales isolated in this study were from non-sterile sites and data were not available regarding clinical symptoms. Although it is presumed these cultures were obtained due to concern for infection at the site of collection, it is possible that the isolated organisms represented colonization. Thus, we were not able to assess the risk of infection alone. Data regarding travel to a foreign country was also not available for analysis so models could not be adjusted for recent travel to a foreign country with high ESBL-E prevalence; however, there was likely less foreign travel during this time due to the COVID-19 pandemic. Further, there may have been missing data due to the retrospective nature of the study that led to misclassification of exposures, although this would not be expected to differ between cases and controls. Additionally, our study only assessed ESBL production in *E. coli, K. pnuemoniae, K. oxytoca*, and *P. mirabilis* in which phenotypic identification of ESBL production was performed routinely by the microbiology lab. Thus, there may have been patients with other ESBL-producing species that were not included. Finally, the prevalence of ESBL-E within each community was estimated based on the proportion of ESBL-E among clinical isolates of *E. coli, K. pneumoniae, K. oxytoca*, and *P. mirabilis* in that community. This may not truly represent the prevalence of ESBL-E in that community given there may be selection bias for which patients had indications for cultures to be obtained.

In summary, our results confirm the importance of several previously identified risk factors for ESBL-E colonization or infection. Additionally, we found a weak association between estimated local prevalence of ESBL-E using CBG aggregations and a patient’s risk of ESBL-E infection or colonization. Further studies are needed to explore whether geographic data can be used to better approximate local communities of transmission, and better capture the relationship between local community prevalence and the risk of infection or colonization.

## Supporting information

Supplementary Information for Census Block Group Clustering Methods

## Data Availability

Mobile phone mobility data from SafeGraph is freely available for research purposes through the SafeGraph COVID-19 Data Consortium. The other data are not publicly available given they are HIPPA protected and were approved for limited use by the Johns Hopkins University School of Medicine Institutional Review Board.

https://www.safegraph.com/covid-19-data-consortium

## Funding Support

This study was supported by the Modeling Infectious Diseases in Healthcare Network (award U01CK000589) and by the Centers for Disease Control and Prevention’s Prevention Epicenters Program (grant number U54CK000617-01-00). This work was also supported by the National Institute of Health T32 AI007291 to J.C. The content is solely the responsibility of the authors and does not necessarily represent the official views of the National Institutes of Health.

## Potential conflicts of interest

All authors report no conflicts of interest relevant to this article.

## Notes

### Competing Interest Statement

The authors have declared no competing interest.

### Funding Statement

This study was supported by the Modeling Infectious Diseases in Healthcare Network (awards U01CK000589) and by the Centers for Disease Control and Preventions Prevention Epicenters Program (grant number U54CK000617-01-00). This work was also supported by the National Institute of Health T32 AI007291 to J.C. The content is solely the responsibility of the authors and does not necessarily represent the official views of the National Institutes of Health.

### Author Declarations

The Institutional Review Board of Johns Hopkins University School of Medicine gave ethical approval for this work with a waiver of informed consent.

