## Supplementary Information for Census Block Group Clustering Methods for "Risk Factor Analysis for Extended-Spectrum Beta-Lactamase Producing Enterobacterales Colonization or Infection: Evaluation of a Novel Approach to Assess Local Prevalence as a Risk Factor"

**Methods**

**Data source**

Our community networks were developed using movement data from SafeGraph ([www.safegraph.com](http://www.safegraph.com)), a data company that aggregates anonymized location data from millions of mobile devices to provide insights about movement patterns. SafeGraph collects hourly data of visits to millions of points of interest (POIs) across the US, which include different types of businesses and establishments, such as retail stores, restaurants, schools, healthcare facilities, etc. The visit dataset includes details such as the number of visits aggregated by the home census block groups (CBGs) of the visitors. SafeGraph data have been used in different studies to investigate movement patterns ^1,2^, especially transmission of COVID-19 in communities ^3,4^.

**Community networks**

Using SafeGraph movement data, a set of community networks were constructed to discover the structure and patterns of connectedness in the population.

*1. Data preparation*

As the spatial scope of this study was the Johns Hopkins Health System catchment area within the Maryland-DC region, the data was filtered for the POIs and CBGs within the study area. For privacy reasons, SafeGraph excludes CBGs for which fewer than five cellphone devices are registered. Furthermore, while SafeGraph categorizes POIs into more than 400 categories, to enable cross-category analyses, we aggregated sub-categories and certain top-categories of SafeGraph into the following nine categories: Retail, Parks, Religious, School, Health, Restaurants, Bars, Other, and Unknown. For this study, we excluded the Health POIs as we are interested in movements outside of the healthcare facilities. Overall, we analyzed visits to 112,207 POIs in 5852 CBGs within the Maryland-DC area from January 2019 to March 2021.

*2. Weekly flow matrices*

We used SafeGraph Weekly Places Patterns datasets to calculate the weekly flow between all pairs of CBGs in the study area. This dataset contains weekly number of visits for each POI and the home CBGs of the visitors. Using the CBGs of POIs as destinations and the home CBGs of the visitors as origins, we aggregated the visit counts by CBG and calculated the weekly flow matrices for each of the 8 category of POIs (excluding Health):

$F_{c,w}=\left[ f_{ij}^{c,w} \right]$,

(1)

where $f_{ij}^{c,w}$ is the weekly total number of visitors from CBG $i$ that visited a POI of category $c$ in CBG $j$ during week $w$. The average weekly flow matrix was then calculated as:

$\bar{F}=\left[ \bar{f}_{ij} \right]=\left[ \sum_{c} \sum_{w} f_{ij}^{c,w}/n_{w} \right]$,

(2)

where $n_{w}$ is the number of weeks.

*3. Average interaction matrix*

Having calculated the average weekly flow matrix, we calculated the average normalized flow matrix, as follows:

$\tilde{F}=[\tilde{f}_{ij}]=\left[ \frac{\bar{f}_{ij}}{n_{i}} \right]$,

(3)

where $\tilde{f}_{ij}$ represents the average proportion of the visits that the population of CBG $i$ make to non-healthcare POIs in CBG $j$ in any week ($0\leq\tilde{f}_{ij}\leq1)$, and $n_{i}$ is the number of registered cellphone devices (in SafeGraph system) in CBG $i$, respectively.

Then, we calculated the interaction matrix that gives the average weekly probability of interaction between every pair of CBGs ($p_{ij}$):

$I=\left[ p_{ij} \right]=\left[ \sum_{k\in V} \left( \tilde{f}_{i,k}\times\tilde{f}_{j,k}\times\frac{\bar{f}_{j,k}}{\sum_{s\in V} \bar{f}_{s,k}} \right) \right]$*.*

(4)

where $V$ is the set of all CBGs.

Each component, $p_{ij}$, of the interaction matrix denotes the probability that any random individual from CBGs $i$ would contact a random individual from CBG $j$ (whether inside CBG $i$, $j$, or any other CBG). In Equation (4), the term $\tilde{f}_{i,k}\times\tilde{f}_{j,k}$ denotes the joint probability that two random individuals from CBGs $i$ and $j$ would go to any CBG $k$, and $\frac{\bar{f}_{j,k}}{\sum_{s\in V} \bar{f}_{s,k}}$ denotes the probability of interaction. The probability that an individual from CBG $i$ contacts an individual from CBG$j$ inside CBG $k$ depends on the active population mixing of CBG $k$ (here, active population refers to the population that actively visit POIs in CBG $k$, and it does not refer to the population of CBG $k$). From all the population actively visiting POIs in CBG $k$, $\sum_{s\in V} \bar{f}_{s,k}$, only $\bar{f}_{j,k}$ are from CBG $j$, hence the probability of random interaction with people from CBG $j$ equals $\frac{\bar{f}_{j,k}}{\sum_{s\in V} \bar{f}_{s,k}}$.

It is noteworthy that the interaction matrix is not necessarily symmetric because the probability of interaction depends on the proportion of the visits that the populations of two CBGs make to all other CBGs, i.e.:

$$p_{ij}=\sum_{k\in V} \left( \tilde{f}_{i,k}\times\tilde{f}_{j,k}\times\frac{\bar{f}_{j,k}}{\sum_{s\in V} \bar{f}_{s,k}} \right),$$

$$p_{ji}=\sum_{k\in V} \left( \tilde{f}_{j,k}\times\tilde{f}_{i,k}\times\frac{\bar{f}_{i,k}}{\sum_{s\in V} \bar{f}_{s,k}} \right),$$

$$\frac{\bar{f}_{j,k}}{\sum_{s\in V} \bar{f}_{s,k}}\neq\frac{\bar{f}_{i,k}}{\sum_{s\in V} \bar{f}_{s,k}} \text{for} i\neq j$$

(5)

which implies $p_{ij}\neq p_{ji}$ for $i\neq j$.

*4. Community detection*

For each CBG in the study area, the corresponding row of the interaction matrix indicates the level of connection with other CBGs in terms of the probability of interaction. We applied a set of community detection methods to discover clusters of CBGs in which residents are more likely to interact with each other rather than with people from other clusters.

First, we created the mobility network based on the average CBG interaction matrix, defined as a directed and edge-weighted graph $G=(V,E,W)$, in which vertices ($V$) are the CBGs, edges ($E$) denote CBGs with any positive level of population interaction, and weights ($W$) are the probabilities of interaction between populations of pairs of CBGs. The result was a network with 2692 nodes and 164,556 weighted edges. For detecting community structures, we used the greedy Clauset-Newman-Moore^5^ and the Louvain algorithm,^6^ which are both based on modularity maximization and among the most commonly used community detection algorithms across different datasets. We conducted sensitivity analysis on the resolution parameter, often called gamma ($\gamma$), of these algorithms. Gamma is a hyper parameter that sets an arbitrary tradeoff between intra-group and inter-group edges. Increasing the resolution parameter can result in smaller-sized clusters and more complex structures. We incrementally increased gamma from 1 to 2. In addition, as Louvain is a heuristic method, we repeated the analysis for each value of gamma and extracted a set of 100 possible community structures for the study area.

Given the probabilistic nature of community detection algorithms and their sensitivity to the value of the resolution parameter, and in the absence of ground truth, it is difficult to judge the reliability of the resulted community structures. To incorporate this uncertainty into our analysis, we applied the method of community cores^7^ to identify clusters of CBGs on which the results of the community detection analysis agree most of the time. The community cores methodology aims at finding the tendency (probability) of pairs of nodes to belong to the same cluster across multiple executions of non-deterministic community detection algorithms or either deterministic or probabilistic algorithms with varying hyper parameters.

To identify the community cores, we created an $n\times n$ matrix, where $n$ is the number of nodes (CBGs), denoted by $M=[m_{ij}]$. Each component of this matrix, $m_{ij}$, is the fraction of executions in which CBGs $i$ and $j$ were classified in the same community (note that $m_{ij}=m_{ji}$). Based on matrix $M$, we create a new undirected weighted network $G^{'}=(V,E^{'},W'$), where the weight of the edge $(i,j)$ is $m_{ij}$. Finally, for any given threshold $\alpha\in[0, 1]$, the edges with $m_{ij}<\alpha$ are removed from $G'$ to obtain the so called α-cores subgraph $G_{\alpha}^{'}$, which is non-overlapping clusters of CBGs that appear in at least in a fraction $\alpha$ of all the resulted community structures.

**CBG Communities**

A sample of resulted community structures using the greedy algorithm is shown in Figure S1. As the resolution parameter was increased from 1 to 1.6, the number of detected communities increased from 8 to 13. A summary of the results of the community cores methodology is shown in Figures S2 and S3. As the threshold increased in value, the number of clusters increased such that for thresholds of 50%, 95%, and 99%, the number of clusters were 3, 74, and 179, respectively.

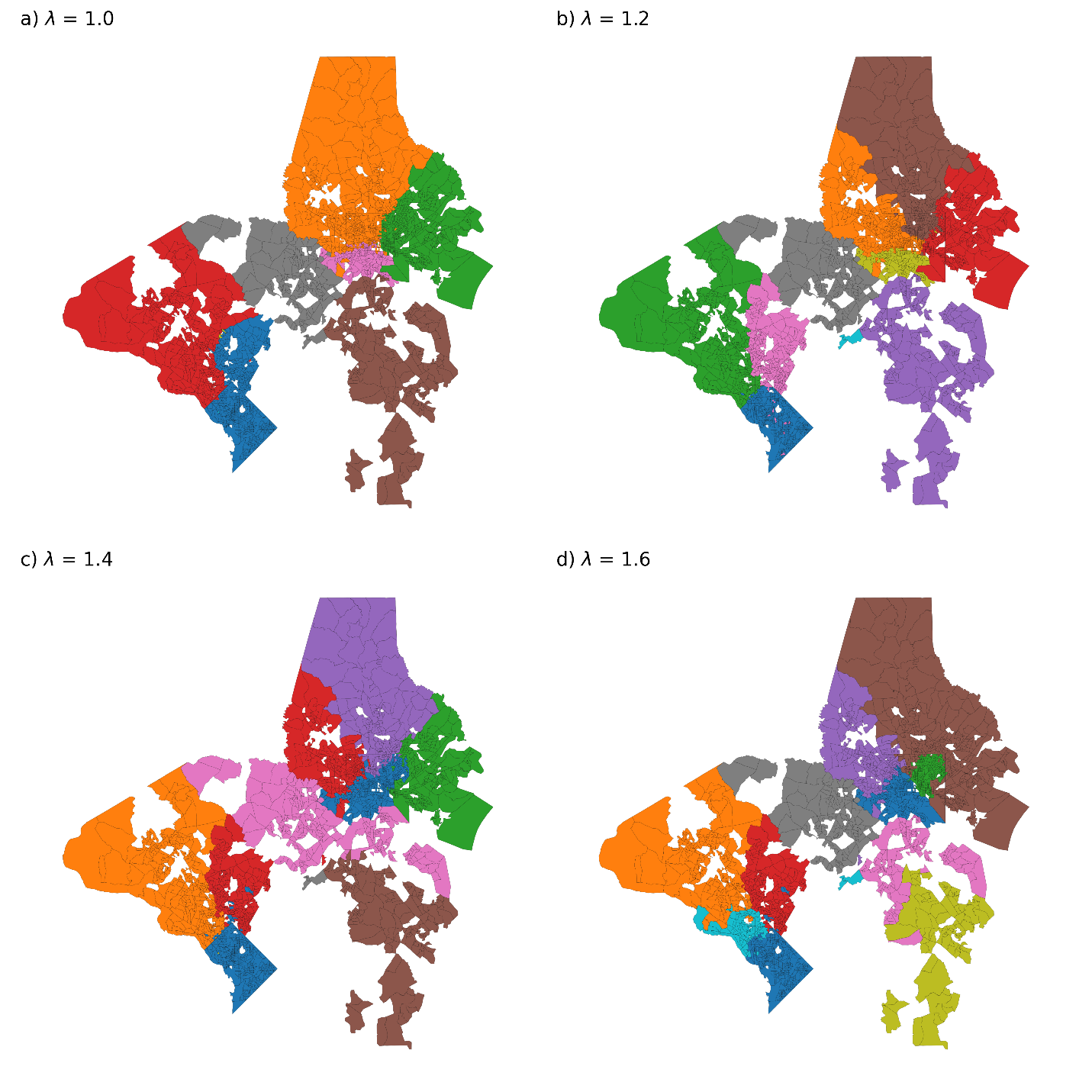

Figure S1. Examples of detected community structures with different resolution parameters.

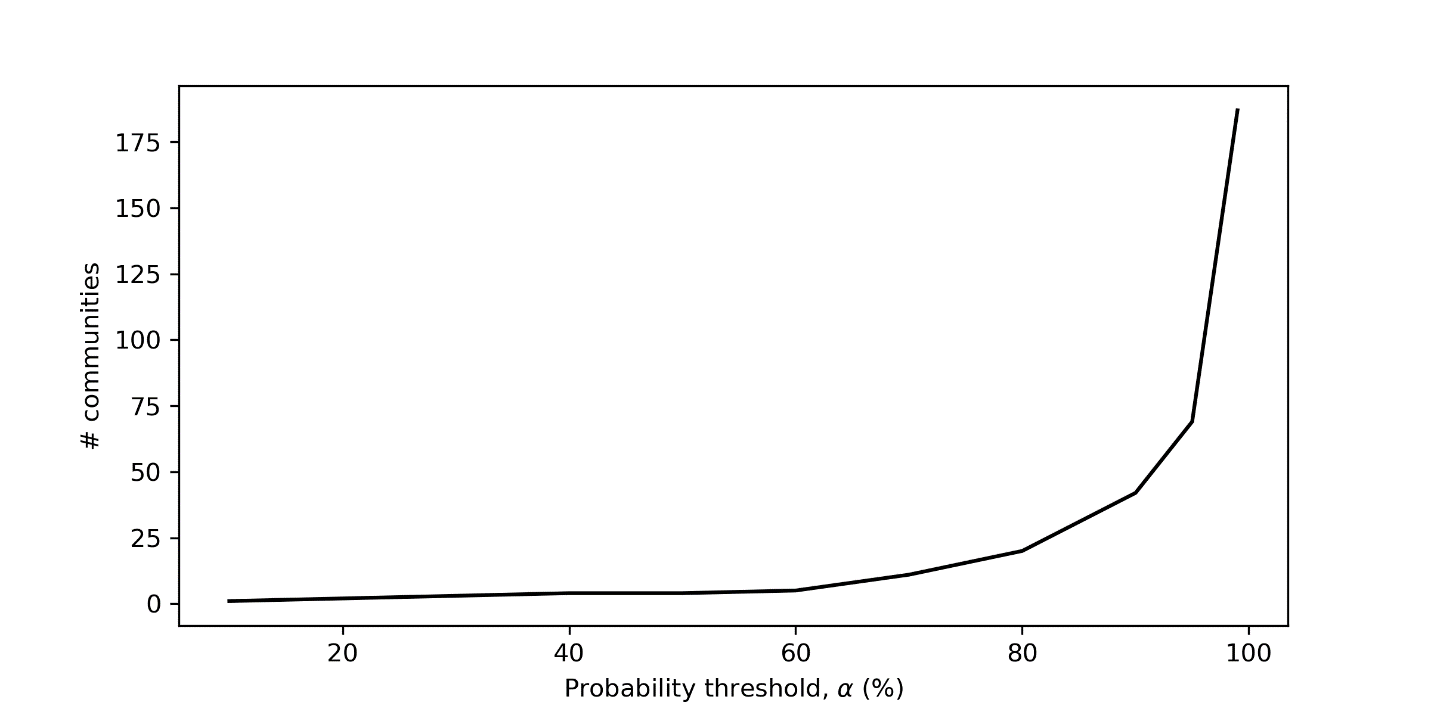

Figure S2. Results of community cores methodology

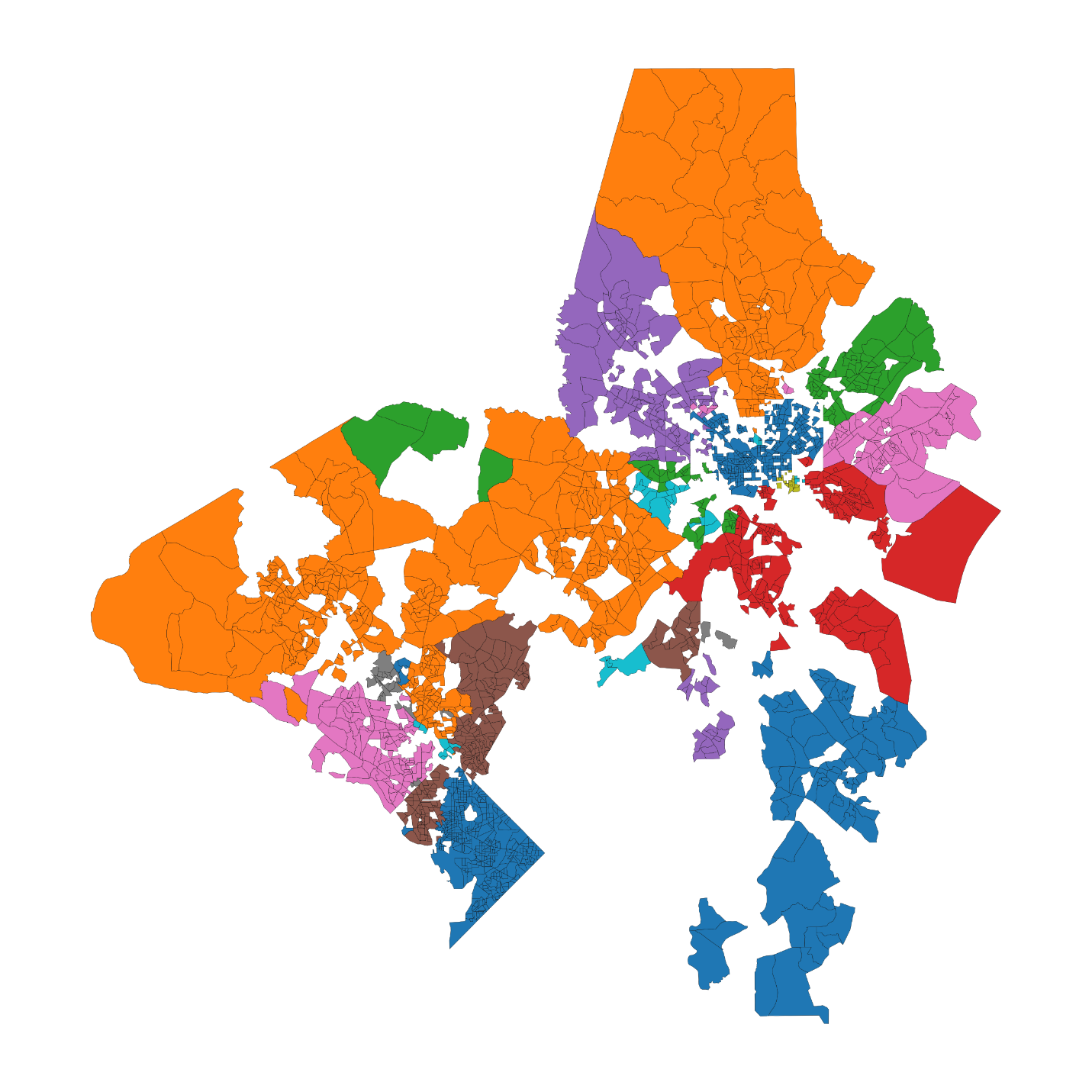

Figure S3. Results of the community cores methodology with a threshold of $\alpha=0.95$.

**Data availability**

Mobile phone mobility data from SafeGraph is freely available for research purposes through the SafeGraph COVID-19 Data Consortium (<https://www.safegraph.com/covid-19-data-consortium>).

**Acknowledgment**

The authors would like to thank SafeGraph for providing their data for COVID-19 related work.
