## Supplementary Table 1 for "Risk Factor Analysis for Extended-Spectrum Beta-Lactamase Producing Enterobacterales Colonization or Infection: Evaluation of a Novel Approach to Assess Local Prevalence as a Risk Factor"

**Table S1.** Univariable Logistic Regression Models Evaluating Risk Factors for Isolation of an Extended-Spectrum β-Lactamase Producing Enterobacterales from Emergency Department Patients

| Baseline Characteristics | Unadjusted Odds Ratio (95% CI) |
| --- | --- |
| Age group |  |
| 18-25 yr | Ref |
| 26-35 yr | 2.18 (1.41-3.38) |
| 36-45 yr | 1.90 (1.21-2.99) |
| 46-55 yr | 2.77 (1.81-4.25) |
| 56-65 yr | 3.19 (2.13-4.80) |
| 66-75 yr | 3.32 (2.23-4.97) |
| 76-85 yr | 2.80 (1.88-4.19) |
| >85 yr | 1.97 (1.30-2.98) |
| Male Sex | 1.61 (1.42-1.82) |
| Race/Ethnicity |  |
| White | Ref |
| Black | 1.21 (1.05-1.40) |
| Asian | 1.60 (1.24-2.06) |
| Hispanic | 1.58 (1.29-1.94) |
| Other | 2.08 (1.58-2.74) |
| Comorbidities |  |
| Diabetes | 1.65 (1.44-1.90) |
| Chronic Kidney Disease | 1.59 (1.36-1.85) |
| Chronic Pulmonary Disease | 1.27 (1.06-1.52) |
| Chronic Liver Disease | 1.03 (0.77-1.40) |
| Cancer | 1.01 (0.85-1.21) |
| HIV Positive | 1.59 (1.33-1.89) |
| Antibiotic Use (<6 Months) |  |
| 3^rd^ Generation Cephalosporin | 3.12 (2.69-3.61) |
| 4^th^ Generation Cephalosporin | 2.77 (2.26-3.38) |
| Piperacillin-Tazobactam | 2.29 (1.86-2.82) |
| Carbapenem | 8.41 (6.77-10.43) |
| Fluoroquinolone | 2.40 (1.87-3.09) |
| Aminoglycoside | 2.28 (1.43-3.62) |
| Trimethoprim-Sulfamethoxazole | 2.99 (2.22-4.04) |
| Aztreonam | 2.67 (1.39-5.11) |
| Metronidazole | 1.84 (1.44-2.36) |
| Other | 1.90 (1.65-2.20) |
| Acid Suppressant Use (<6 Months) |  |
| Proton Pump Inhibitor | 2.15 (1.85-2.51) |
| H2 Antagonist | 1.92 (1.57-2.35) |
| Device or Hardware at Presentation |  |
| Urinary Catheter | 2.35 (1.78-3.11) |
| Tracheostomy | 8.13 (3.92-16.89) |
| Gastrointestinal Feeding Tube | 2.44 (1.16-5.12) |
| Central Line | 1.64 (1.12-2.41) |
| Healthcare Exposure |  |
| Hospitalization (<12 months) | 2.09 (1.84-2.38) |
| ICU Admission (<12 months) | 2.63 (2.13-3.25) |
| Surgery or Procedure (<12 months) | 1.63 (1.42-1.88) |
| Long Term Care Facility or Skilled Nursing Facility Admission (<6 Months) | 2.20 (1.90-2.54) |
| History of ESBL-E (<12 months) | 28.76 (21.64-38.23) |
| Prevalence of ESBL-E within Patient’s CBG Community (<3 months) |  |
| <25^th^ Percentile | 0.84 (0.72-0.98) |
| 25^th^ to 75^th^ Percentile | Ref |
| >75^th^ Percentile | 0.93 (0.80-1.08) |
| Prevalence of ESBL-E within Patient’s CBG Community (<6 months) |  |
| <25^th^ Percentile | 0.86 (0.74-1.01) |
| 25^th^ to 50^th^ Percentile | Ref |
| >75^th^ Percentile | 0.91 (0.79-1.06) |
| Prevalence of ESBL-E within Patient’s CBG Community (<12 months) |  |
| <25^th^ Percentile | 0.84 (0.72-0.98) |
| 25^th^ to 50^th^ Percentile | Ref |
| >75^th^ Percentile | 0.99 (0.85-1.14) |

Abbreviations: ESBL-E = Extended-spectrum β-lactamase producing Enterobacterales; CBG = Census block group; ICU = Intensive care unit; H-2 = Histamine H2-receptor; HIV = Human Immunodeficiency Virus
