## Supplementary Table 2 for "Risk Factor Analysis for Extended-Spectrum Beta-Lactamase Producing Enterobacterales Colonization or Infection: Evaluation of a Novel Approach to Assess Local Prevalence as a Risk Factor"

**Table S2.** Multivariable Logistic Regression Models Evaluating Risk Factors for Isolation of an Extended-Spectrum β-Lactamase (ESBL) Producing Enterobacterales from Emergency Department Patients without an ESBL Positive Culture in the Past Year

| Baseline Characteristics | Model 1^*^ | Model 2^†^ | Model 3^ǂ^ | Model 4^¶^ |
| --- | --- | --- | --- | --- |
|  | Adjusted OR (95% CI) | | | |
| Age group |  |  |  |  |
| 18-25 yr | Ref | Ref | Ref | Ref |
| 26-35 yr | 2.21 (1.37-3.56) | 2.21 (1.37-3.56) | 2.20 (1.37-3.57) | 2.20 (1.36-3.55) |
| 36-45 yr | 1.80 (1.10-2.95) | 1.81 (1.10-2.96) | 1.80 (1.10-2.96) | 1.80 (1.10-2.94) |
| 46-55 yr | 2.53 (1.58-4.06) | 2.54 (1.58-4.07) | 2.54 (1.58-4.07) | 2.52 (1.57-4.05) |
| 56-65 yr | 3.06 (1.94-4.82) | 3.06 (1.94-4.82) | 3.06 (1.94-4.82) | 3.05 (1.94-4.81) |
| 66-75 yr | 3.13 (1.99-4.91) | 3.15 (2.00-4.94) | 3.14 (2.00-4.94) | 3.13 (2.00-4.91) |
| 76-85 yr | 2.52 (1.60-3.96) | 2.52 (1.60-3.97) | 2.52 (1.60-3.97) | 2.52 (1.60-3.96) |
| >85 yr | 1.82 (1.14-2.91) | 1.82 (1.14-2.92) | 1.82 (1.14-2.91) | 1.81 (1.13-2.89) |
| Male Sex | 1.29 (1.12-1.50) | 1.30 (1.12-1.49) | 1.29 (1.12-1.50) | 1.29 (1.12-1.50) |
| Race/Ethnicity |  |  |  |  |
| White | Ref | Ref | Ref | Ref |
| Black | 1.26 (1.07-1.49) | 1.26 (1.07-1.48) | 1.26 (1.07-1.48) | 1.25 (1.06-1.47) |
| Asian | 1.85 (1.40-2.45) | 1.83 (1.38-2.43) | 1.84 (1.39-2.44) | 1.83 (1.38-2.42) |
| Hispanic | 2.31 (1.83-2.92) | 2.32 (1.84-2.92) | 2.32 (1.83-2.92) | 2.29 (1.82-2.90) |
| Other | 2.65 (1.95-3.59) | 2.62 (1.94-3.56) | 2.63 (1.94-3.57) | 2.62 (1.94-3.56) |
| Comorbidities |  |  |  |  |
| Diabetes | 1.18 (1.00-1.40) | 1.18 (1.00-1.40) | 1.18 (1.00-1.40) | 1.18 (1.00-1.40) |
| Chronic Kidney Disease | 1.15 (0.95-1.40) | 1.15 (0.95-1.40) | 1.15 (0.95-1.40) | 1.16 (0.96-1.40) |
| Chronic Pulmonary Disease | 1.00 (0.80-1.25) | 1.00 (0.81-1.25) | 1.00 (0.81-1.25) | 1.01 (0.81-1.26) |
| Chronic Liver Disease | 0.70 (0.50-1.00) | 0.70 (0.50-1.00) | 0.70 (0.50-1.00) | 0.70 (0.50-1.00) |
| Cancer | 0.78 (0.63-0.96) | 0.78 (0.63-0.96) | 0.78 (0.62-0.96) | 0.77 (0.63-0.95) |
| HIV Positive | 1.24 (1.00-1.53) | 1.24 (1.00-1.54) | 1.25 (1.00-1.54) | 1.25 (1.00-1.55) |
| Antibiotic Use (<6 Months) |  |  |  |  |
| 3^rd^ Generation Cephalosporin | 1.82 (1.47-2.25) | 1.83 (1.48-2.26) | 1.83 (1.48-2.27) | 1.83 (1.48-2.27) |
| 4^th^ Generation Cephalosporin | 1.08 (0.80-1.46) | 1.08 (0.80-1.47) | 1.08 (0.80-1.46) | 1.09 (0.81-1.48) |
| Piperacillin-Tazobactam | 0.81 (0.60-1.10) | 0.81 (0.60-1.10) | 0.81 (0.60-1.10) | 0.81 (0.60-1.09) |
| Carbapenem | 2.64 (1.85-3.77) | 2.62 (1.83-3.74) | 2.64 (1.84-3.77) | 2.64 (1.85-3.78) |
| Fluoroquinolone | 1.27 (0.91-1.78) | 1.27 (0.91-1.78) | 1.27 (0.91-1.78) | 1.28 (0.91-1.79) |
| Aminoglycoside | 0.80 (0.39-1.64) | 0.80 (0.39-1.64) | 0.78 (0.39-1.64) | 0.80 (0.39-1.64) |
| Trimethoprim-Sulfamethoxazole | 1.66 (1.13-2.44) | 1.66 (1.13-2.44) | 1.65 (1.12-2.43) | 1.65 (1.12-2.44) |
| Aztreonam | 1.09 (0.49-2.43) | 1.08 (0.48-2.40) | 1.08 (0.49-2.41) | 1.08 (0.49-2.41) |
| Metronidazole | 0.89 (0.63-1.25) | 0.88 (0.63-1.25) | 0.89 (0.63-1.25) | 0.89 (0.63-1.26) |
| Other | 0.95 (0.77-1.17) | 0.94 (0.77-1.16) | 0.95 (0.77-1.16) | 0.95 (0.77-1.16) |
| Acid Suppressant Use (<6 Months) |  |  |  |  |
| Proton Pump Inhibitor | 0.93 (0.74-1.17) | 0.93 (0.74-1.17) | 0.93 (0.74-1.17) | 0.93 (0.74-1.17) |
| H2 Antagonist | 1.11 (0.85-1.45) | 1.11 (0.85-1.45) | 1.11 (0.84-1.45) | 1.09 (0.83-1.43) |
| Device or Hardware at Presentation |  |  |  |  |
| Urinary Catheter | 1.36 (0.97-1.91) | 1.37 (0.97-1.92) | 1.37 (0.98-1.93) | 1.36 (0.97-1.91) |
| Tracheostomy | 3.60 (1.50-8.62) | 3.55 (1.48-8.48) | 3.56 (1.49-8.52) | 3.53 (1.47-8.49) |
| Gastrointestinal Feeding Tube | 0.91 (0.35-2.33) | 0.89 (0.35-2.30) | 0.91 (0.35-2.32) | 0.92 (0.36-2.35) |
| Central Line | 0.94 (0.59-1.50) | 0.93 (0.58-1.48) | 0.92 (0.58-1.47) | 0.92 (0.58-1.47) |
| Healthcare Exposure |  |  |  |  |
| Hospitalization (<12 months) | 0.89 (0.73-1.09) | 0.89 (0.73-1.09) | 0.89 (0.73-1.09) | 0.89 (0.73-1.09) |
| ICU Admission (<12 months) | 1.25 (0.92-1.69) | 1.25 (0.93-1.70) | 1.26 (0.93-1.71) | 1.26 (0.93-1.70) |
| Surgery or Procedure (<12 months) | 1.13 (0.95-1.36) | 1.14 (0.95-1.36) | 1.14 (0.95-1.36) | 1.13 (0.95-1.36) |
| Long Term Care Facility or Skilled Nursing Facility Admission (<6 Months) | 1.73 (1.45-2.09) | 1.73 (1.44-2.09) | 1.72 (1.44-2.08) | 1.73 (1.44-2.09) |
| Prevalence of ESBL-E within Patient’s CBG Community Aggregate |  |  |  |  |
| <25^th^ Percentile | - | 0.82 (0.69-0.97) | 0.83 (0.70-0.98) | 0.78 (0.66-0.93) |
| 25^th^ to 75^th^ Percentile | - | Ref | Ref | Ref |
| >75^th^ Percentile | - | 0.96 (0.82-1.13) | 0.95 (0.81-1.12) | 1.01 (0.85-1.18) |

Abbreviations: CBG = Census block group; ESBL-E = Extended-spectrum β-lactamase producing Enterobacterales; H-2 = Histamine H2-receptor; HIV = Human Immunodeficiency Virus; ICU = Intensive care unit; OR = Odds Ratio

^*^Model 1: Includes previously identified risk factors in the literature as dichotomous variables.

^†^Model 2: Model 1 and a categorical variable indicating percentile for proportion of ESBL-producing isolates within the patient’s CBG community in the previous 3 months.

^ǂ^Model 3: Model 1 and a categorical variable indicating percentile for proportion of ESBL-producing isolates within the patient’s CBG community in the previous 6 months.

^¶^Model 4: Model 1 and a categorical variable indicating percentile for proportion of ESBL-producing isolates within the patient’s CBG community in the previous 12 months.
